# Neuron-Specific DNA Methylation Differences in the Prefrontal Cortex in Parkinson’s Disease

**DOI:** 10.64898/2026.02.03.26344617

**Authors:** Anthony Klokkaris, Eilis Hannon, Joe Burrage, Barry Chioza, Adam R. Smith, Joshua Harvey, Alice Franklin, Luke Weymouth, Jennifer Imm, Katie Lunnon, Emma L. Dempster, Jonathan Mill, Anna Migdalska-Richards

## Abstract

Parkinson’s disease (PD) is a progressive movement disorder that affects over ten million individuals worldwide. While the involvement of genetically-driven cellular mechanisms in PD pathogenesis is well-established, there is increasing evidence that epigenetic dysregulation also plays a key role.

We profiled genome-wide DNA methylation in isolated neuronal, oligodendrocyte and other glial nuclei populations from the prefrontal cortex of 71 PD and 56 control individuals. We identified seven significant differentially methylated positions in neuronal nuclei associated with PD. All these sites were hypermethylated in PD, with five of the differentially methylated positions located in the following genes: *ROBO4, SSBP2, PDE4B, NPHP1*, and *HSD17B12*. No differentially methylated positions were observed in oligodendrocyte or other glial nuclei, highlighting the neuronal specificity of PD-associated methylation changes. Comparison with a large bulk brain meta-analysis of Lewy body pathology confirmed concordant directionality for ∼79% of neuronal differentially methylated positions, indicating that bulk tissue signals primarily reflect neuronal alterations.

Together, these findings provide the first cell type-resolved map of DNA methylation changes in the PD cortex, revealing neuronal-specific hypermethylation at novel loci and emphasizing the importance of cell type-specific analyses in disentangling the molecular heterogeneity of PD. This study lays the groundwork for future multi-omics and region-specific studies aimed at uncovering mechanisms underlying disease vulnerability and progression at the cellular level.

## Introduction

Parkinson’s disease (PD) is a progressive movement disorder and the second most prevalent cause of neurodegeneration, affecting over ten million people globally [1]. The incidence of PD increases markedly with advancing age, with approximately 1.7% of individuals aged 80–84 affected [2]. Epidemiological projections indicate that the prevalence of PD will double between 2015 and 2040 [3]. Pathologically, PD is characterized by the progressive loss of dopaminergic neurons in the substantia nigra, which is normally accompanied by the formation of α-synuclein aggregates in intracellular cytoplasmic Lewy body (LB) inclusions – the pathological hallmark of PD [4]. Despite considerable advances in research, there is still a lack of understanding of the etiological mechanisms underlying PD, and no disease-modifying therapies are currently available.

A small proportion of PD cases can be attributed to rare, highly penetrant mutations in specific genes, including α-synuclein (*SNCA*), leucine-rich repeat kinase 2 (*LRRK2*), PTEN-induced kinase 1 (*PINK1*), parkin (*PRKN*), parkinsonism-associated deglycase (*DJ-1*) and ATPase cation transporting 13A2 (*ATP13A2*) [5]. Moreover, several common genetic variants have been identified as risk factors for idiopathic PD, including allelic variants within the glucocerebrosidase 1 (*GBA1*) gene [6]. *GBA1* encodes the lysosomal enzyme β-glucocerebrosidase which is essential for sphingolipid metabolism through the breakdown of glucosylceramide and glucosylsphingosine [6]. *GBA1* mutations represent the most common genetic risk factor for nonfamilial PD [7], with an estimated 5–10% prevalence among non-Ashkenazi Jewish PD individuals [7–9].

Genome-wide association studies (GWAS) have further expanded the number of common genetic loci known to be associated with PD; a 2019 meta-analysis of 17 GWAS reported 90 independent risk signals [10], with additional loci of smaller effect likely to be uncovered as even larger meta-analyses are undertaken in the field. However, as most GWAS variants map to non-coding regulatory regions [11] and monozygotic twin concordance for PD is only ∼17% [12], non-genetic factors, including epigenetic variation, are likely to contribute substantially to disease pathogenesis.

DNA methylation is the most extensively studied epigenetic modification, involving the addition of a methyl group to the 5-carbon cytosine to form 5-methylcytosine [13,14]. It is well established that DNA methylation is associated with aging [15–17], the primary risk factor for PD. Altered DNA methylation patterns have been increasingly implicated in PD and other neurological and neuropsychiatric disorders such as Alzheimer’s disease (AD) and schizophrenia [18–21].

Early DNA methylation studies in PD primarily focused on individual genes previously implicated in pathogenesis, most notably *SNCA*, where intron 1 hypomethylation has been consistently reported in PD brains [22–27]. Since then, multiple epigenome-wide association studies (EWAS) have identified several differentially methylated positions (DMPs) in PD, including in the frontal cortex and substantia nigra [28–35]. Some studies have also looked at DNA methylation in relation to LB pathology. An EWAS by Pihlstrøm et al. (2022) that was undertaken in frontal cortex tissue from PD and dementia with Lewy body (DLB) donors identified 24 DMPs associated with Braak LB stage, a measure of LB spread through the brain [36]. More recently, a large meta-analysis of DNA methylation in the cortex reported DMPs associated with Braak LB stage [37].

A key limitation of most PD epigenetic studies is their use of bulk brain tissue, which reflects DNA methylation averaged across heterogenous cell types including neurons, oligodendrocytes, microglia and astrocytes – each known to contribute differently to PD pathogenesis [48–52]. Given that DNA methylation profiles are highly cell type-specific [21,43,44], a few studies in PD have applied approaches to isolate neuronal nuclei from cortical tissue for DNA methylation profiling [45–50]. For instance, Marshall et al. identified 1,799 PD-associated DMPs after false-discovery rate (FDR) correction in enhancers in neuronal nuclei isolated from the prefrontal cortex [46]. These were predicted to regulate genes including *DJ-1, PRKN* and *TET2* – a key enzyme in the demethylation pathway. Additionally, Kochmanski et al. reported 90 sex-specific PD-associated DMPs (P < 9.0×10^-08^) in neuronal nuclei from the parietal cortex [50]. Another study examined DNA methylation at *SNCA* intron 1 separately in neuronal and glial nuclei compared to bulk tissue from the frontal cortex [48]. They observed lower DNA methylation levels in PD compared to controls only in neuronal nuclei.

One study has stratified individuals by *GBA1* status [51], utilizing bisulfite pyrosequencing to assess DNA methylation in intron 1 of the *SNCA* gene in PD-*GBA1* and idiopathic PD cases. They reported hypomethylation at several CpG sites in the PD-*GBA1* group only [51]. Additional cell type separation and genetic stratification in a genome-wide context may yield a more nuanced understanding of DNA methylation alterations in PD.

In this study, we present a comprehensive brain cell specific EWAS of PD. For the first time, we simultaneously profiled genome-wide DNA methylation in isolated prefrontal cortex nuclei populations enriched for neurons, oligodendrocytes, and other glial cells from PD and control donors with *GBA1* genotyping. Here, we leveraged a fluorescence-activated nuclei sorting (FANS) protocol developed by our team [52,53]. Our key finding was the identification of seven genome-wide significant DMPs associated with PD status in neuronal nuclei, highlighting neuronal-specific DNA methylation differences in PD.

## Results

### Study overview

Prefrontal cortex tissue was obtained from a total of 71 PD and 56 control individuals, which were subjected to FANS to isolate nuclei populations enriched for neurons (NeuN+), oligodendrocytes (NeuN-/SOX10+, referred to as SOX10+) and other glia (NeuN-/SOX10-or double negative), with subsequent methylomic profiling on the Infinium MethylationEPIC v1.0 array. Following all quality control (QC) steps and filtering based on estimated cellular purity using the cell type deconvolution goodness (CETYGO) algorithm [54,55], 117 NeuN+ (65 PD and 52 control), 92 SOX10+ (50 PD and 42 control) and 105 double negative (60 PD and 45 control) samples and 736,078 DNA methylation sites remained for EWAS analyses. A breakdown of sample numbers by both PD and *GBA1* status (**Supplementary Data 1**) is provided along with sample-level QC and estimated cellular proportion metrics (**Supplementary Data 2-3**). Principal component analysis (PCA) on the final filtered and normalized dataset demonstrated that cell types distinctly cluster on a graph of the first two principal components (PCs) which explain most of the data variation (PC1 = 57.2%, PC2 = 26.9%) (**Supplementary Figure 1**). We performed power calculations for each cell type separately based on the smallest group size out of PD and controls (see Materials and Methods). The mean differences in DNA methylation (%) in each cell type that could be detected in 80% of CpG sites with at least 80% power for NeuN+, SOX10+ and double negative were 4.9%, 5.7% and 8.8%, respectively (**Supplementary Figure 2**).

PD and control groups did not significantly differ in age, post-mortem interval (PMI), sex ratio (**Table 1**) or cell composition estimates (P > 0.05) (**Supplementary Data 4, Supplementary Figure 3**). The most frequent *GBA1* variant in both cases and controls was E326K. All PD cases had a LB Braak stage > 4, whilst all control samples had a LB Braak stage of 0. Given the availability of post-mortem samples, there were some cases with coincidental AD pathology across both the PD and control groups, however no sample exceeded a Braak neurofibrillary tangle (NFT) stage of IV. There was a significantly higher mean Braak NFT stage in the PD group compared to the control group (P = 2.95×10^-06^, mean difference of 0.6) (**Table 1**) as well as NFT stage differences between certain subgroups when samples were divided by PD and *GBA1* status (Tukey HSD adjusted P < 0.05) (**Supplementary Data 5**).

**Table 1.**
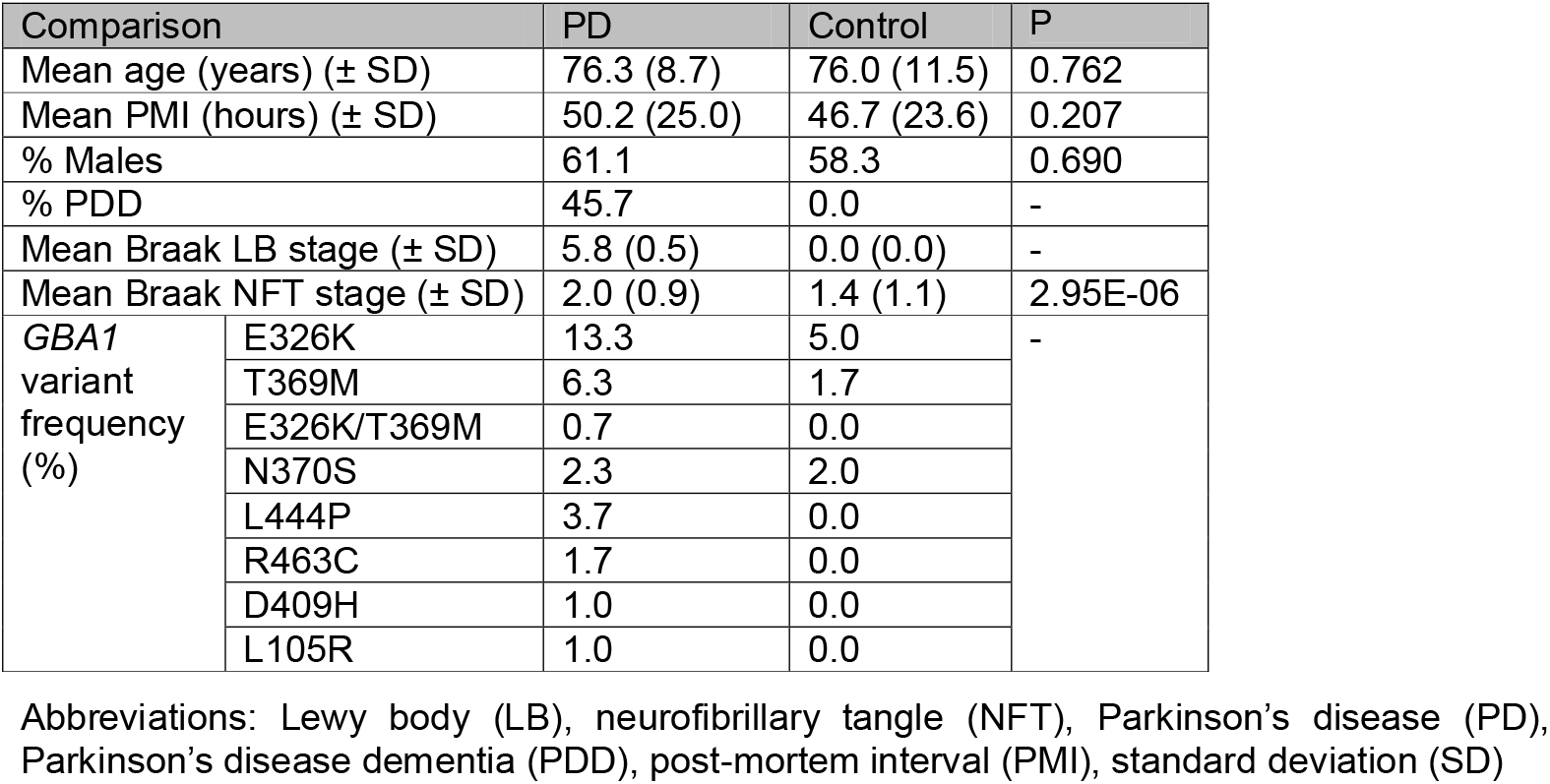
Comparison of sample demographics for the PD and control groups. Sample demographics between the PD and control groups, considering samples in all cell types that passed QC. P values were calculated using a two-sample t-test (age, PMI, Braak NFT stage) or χ^2^ test (sex). 78 NeuN+, 63 SOX10+ and 70 double negative samples had Braak LB stage available, while 95 NeuN+, 74 SOX10+ and 85 double negative samples had Braak NFT stage available.

### Cell type-specific methylomic profiling highlights seven genome-wide significant loci in PD neuronal nuclei

A linear regression analysis was performed in each cell type to identify DMPs associated with PD status after adjusting for confounding variables (**Supplementary Data 6-8**) (see Materials and Methods). λ values showed no inflation for NeuN+ (λ = 0.96) and double negative (λ = 0.96) but mild inflation for SOX10+ (λ = 1.23) (**Figure 1Figure** 1. To identify genome-wide significant DMPs, an empirically-derived significance threshold of P < 9.0×10^-08^ was used [56]. Seven DMPs were associated with PD status in the analysis of NeuN+ samples, all of which were hypermethylated in PD (**Table 2, Figure 1b**). These were cg18248499 (annotated to *ROBO4*, estimate of mean difference in beta values between groups = 0.049, P = 4.02E-10), cg09320924 (annotated to *SSBP2*, estimate = 0.049, P = 8.64E-10), cg27107882 (estimate = 0.039, P = 9.03×10^-09^), cg13471188 (estimate = 0.045, P = 1.06×10^-08^), cg00736744 (annotated to *PDE4B*, estimate = 0.048, P = 2.29×10^-08^), cg11235594 (annotated to *NPHP1*, estimate = 0.026, P = 2.49×10^-08^) and cg14489013 (annotated to *HSD17B12*, estimate = 0.025, P = 5.15×10^-08^). The DMPs were annotated to the gene bodies of *SSBP2, PDE4B, NPHP1* and *HSD17B12*, and upstream of the transcription start site of *ROBO4*.

**Figure 1:**
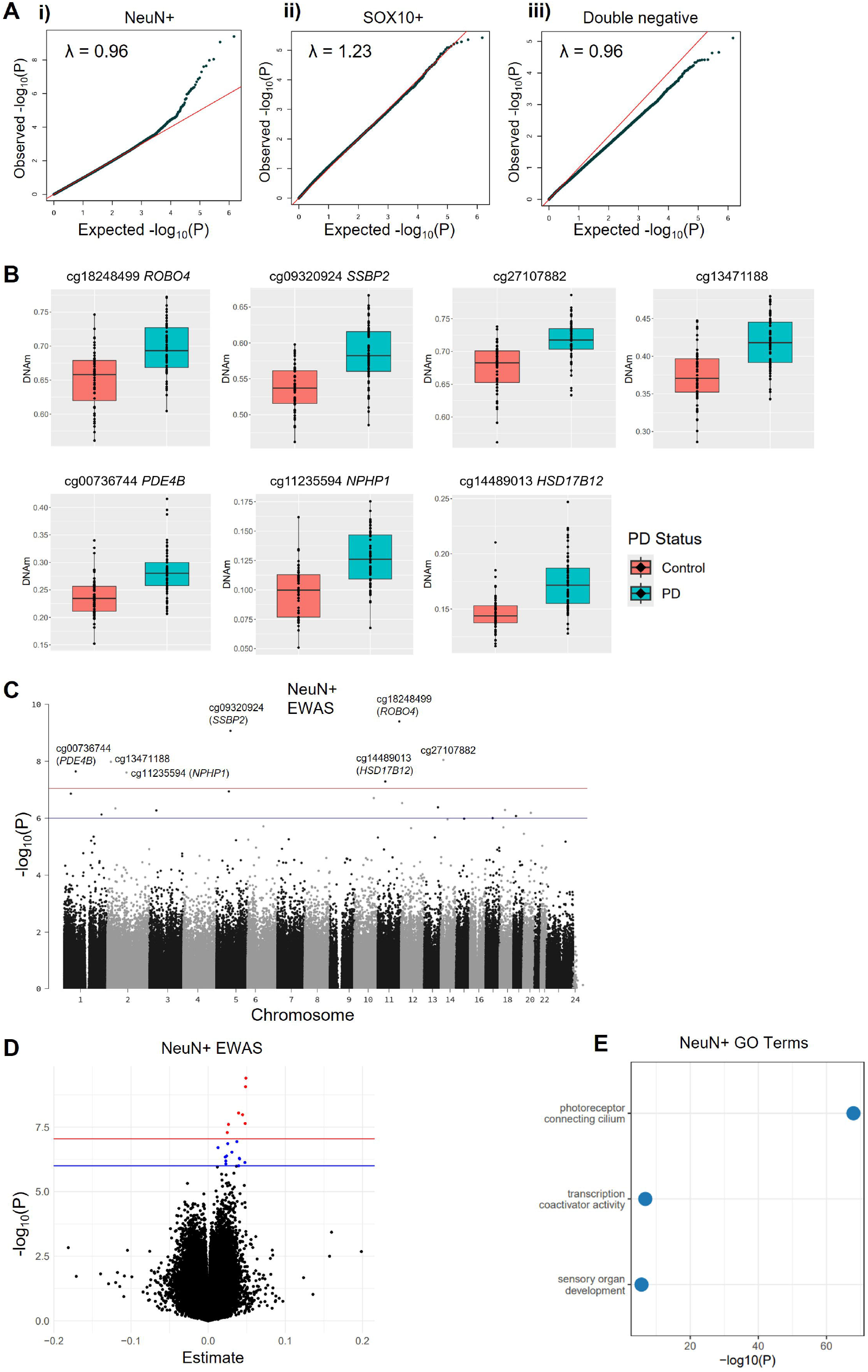
Identification of PD-associated DMPs. (**A**) Quantile-quantile plots for EWASs within each cell type, showing the correlation between the expected -log10(P) values and the observed -log10(P) values for the EWAS with (**i**) NeuN+, (**ii**) SOX10+ and (**iii**) double negative samples. The red line denotes a theoretical, completely linear relationship (y=x). The black dots correspond to the observed p-values for each methylation site included in the analysis. (**B**) Distribution of DNA methylation values for the seven NeuN+ genome-wide significant PD-associated DMPs (P < 9.0×10^-08^). Each plot shows the distribution of normalized DNA methylation beta values in control NeuN+ and PD NeuN+ samples, where each point denotes an individual. Plots are labelled according to the DNA methylation site, along with the nearest gene (Illumina UCSC gene annotation), if a DMP was annotated to any gene. (**C**) Manhattan plots for the EWAS with NeuN+ samples. The X axis shows chromosome number, and the Y axis shows -log10(P). The red line and blue line represent the genome-wide (P = 9.0×10^-08^) and suggestive (P = 1.0×10^-06^) significance thresholds, respectively, with genome-wide significant DMPs labelled along with any annotated gene names in brackets. (**D**) Volcano plots of DNA methylation sites for the EWAS with the NeuN+ samples. The X-axis shows the estimate of the mean difference between the PD and control groups and the Y-axis shows -log10(P). The red line and blue line represent the genome-wide (P = 9.0×10^-08^) and suggestive (P = 1.0×10^-06^) significance thresholds, respectively. Each dot is an individual DNA methylation site, coloured in red if they are genome-wide significant, or blue if suggestively significant. (**E**) -log10(P) for significant gene ontology terms (after Bonferroni correction for the number of pathways tested) from the genes annotated to the genome-wide and suggestively significant NeuN+ DMPs. Abbreviations: epigenome-wide association study (EWAS), gene ontology (GO)

**Table 2:**
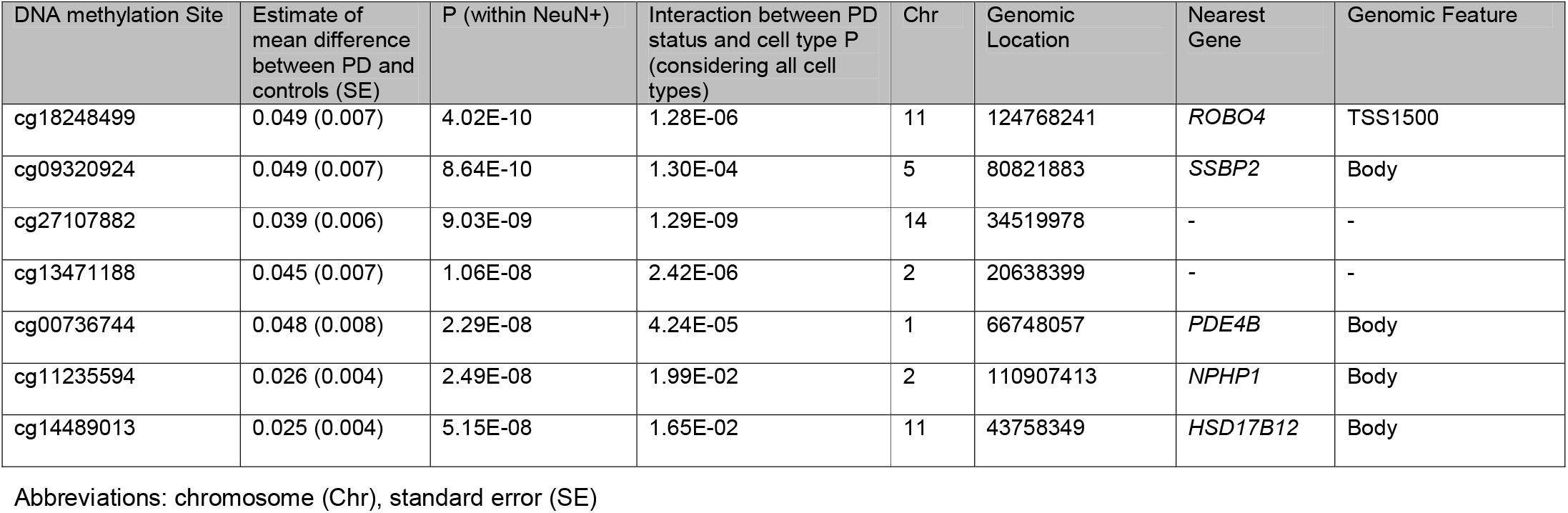
DMPs associated with PD in NeuN+ nuclei at a genome-wide significance threshold (P < 9.0×10^-08^). Seven DMPs reached genome-wide significance for their association with PD in the EWAS with NeuN? samples (n= 65 PD and 52 controls). The estimate refers to the mean difference between the PD and control groups (where DNA methylation is measured as a proportion). P (within NeuN+) refers to the P value from the EWAS with NeuN+ samples only. The interaction between PD status and cell type P column shows the output from the cell type specificity assessment (considering all cell types) using a mixed effects model. Here, an interaction P value < 0.05 indicates that the DMP is cell type-specific. The chromosome number and genomic location of each probe is provided. Probes were annotated to genes using the UCSC gene annotation (based on h19/GRCh37 annotation).

| DNA methylation Site | Estimate of mean difference between PD and controls (SE) | P (within NeuN+) | Interaction between PD status and cell type P (considering all cell types) | Chr | Genomic Location | Nearest Gene | Genomic Feature |
| --- | --- | --- | --- | --- | --- | --- | --- |
| cg18248499 | 0.049 (0.007) | 4.02E-10 | 1.28E-06 | 11 | 124768241 | *ROBO4* | TSS1500 |
| cg09320924 | 0.049 (0.007) | 8.64E-10 | 1.30E-04 | 5 | 80821883 | *SSBP2* | Body |
| cg27107882 | 0.039 (0.006) | 9.03E-09 | 1.29E-09 | 14 | 34519978 | - | - |
| cg13471188 | 0.045 (0.007) | 1.06E-08 | 2.42E-06 | 2 | 20638399 | - | - |
| cg00736744 | 0.048 (0.008) | 2.29E-08 | 4.24E-05 | 1 | 66748057 | *PDE4B* | Body |
| cg11235594 | 0.026 (0.004) | 2.49E-08 | 1.99E-02 | 2 | 110907413 | *NPHP1* | Body |
| cg14489013 | 0.025 (0.004) | 5.15E-08 | 1.65E-02 | 11 | 43758349 | *HSD17B12* | Body |

Abbreviations: chromosome (Chr), standard error (SE)

To further characterize DNA methylation differences, a more lenient suggestive significance threshold of P < 1.0×10^-06^ was applied. Here, an additional 12 DMPs were associated with PD status in the NeuN+ samples which were also all hypermethylated in the disease, giving a total of 19 DMPs at the suggestive significance threshold (**Supplementary Data 9, Figure 1c-d, Supplementary Figure 4**). No DNA methylation sites were associated with PD status in the analysis of SOX10+ or double negative samples at even the suggestive significance threshold (P < 1.0×10^-06^). This highlights that the λ value inflation in the EWAS with SOX10+ samples did not lead to systematic problems. Gene ontology (GO) analysis was performed to identify whether any of the genes annotated to the 19 NeuN+ (P < 1.0×10^-06^) DMPs were enriched in specific pathways (see Materials and Methods). After Bonferroni correction for the number of pathways tested, three significant GO terms were identified (**Figure 1e, Supplementary Data 10**) – photoreceptor connecting cilium (GO:0032391, P=1.20×10^-68^), transcription coactivator activity (GO:0003713, P= 1.62×10^-07^) and sensory organ development (GO:0007423, P= 2.13×10^-06^).

Next, the cell type-specificity of all genome-wide and suggestively significant NeuN+ DMPs was assessed using a mixed effects model (see Materials and Methods). The P-value for the cell type and PD status interaction term was < 0.05 for all DMPs (**Table 2, Supplementary Data 9**) indicating that, for these sites, there is heterogeneity in the PD effects across the cell types and that the difference is likely to be specific to neuronal nuclei.

Given that the PD group had a higher Braak NFT stage than the control group, we performed a sensitivity analysis – repeating the EWAS with NeuN+ samples, with Braak NFT stage as an additional covariate. Of our seven genome-wide significant DMPs, three remained genome-wide significant (cg09320924, cg18248499, cg00736744), while the other four narrowly missed the threshold but remained suggestively significant (cg27107882, cg13471188, cg11235594, cg14489013, all p ≤ 2.82×10^-07^) (**Supplementary Data 11**). For all original NeuN+ DMPs, effects were highly correlated between the original EWAS and sensitivity analysis (Pearson correlation coefficient = 0.968) (**Supplementary Figure 5**) and all directions were consistent, indicating a minimal effect of Braak NFT stage on the main results.

Following this, we examined whether DNA methylation profiles in PD differ according to *GBA1* variant status in each cell type (**Supplementary Data 12-14**; see Materials and Methods). While one probe in NeuN+ samples (cg00727334) was significantly associated with an interaction between *GBA1* status and PD status, visual inspection of this result showed it was due to outliers (**Supplementary Figure 6**), highlighting that we likely did not have enough samples to explore this more thoroughly.

### Validation of PD-associated neuronal DMPs using bulk brain methylation meta-analysis

We compared the PD-associated DMPs in this study with a meta-analysis of DNA methylation in the prefrontal and anterior cingulate cortex in relation to LB pathology burden, which included both PD and DLB cases [37]. Taking their summary statistics corresponding to our seven PD-associated NeuN+ DMPs, we saw that three of these sites showed a significant difference in DNA methylation in the meta-analysis after correcting for seven tests (P < 7.14×10^-03^), with those being cg09320924 (*SSBP2*, P=2.00×10^-03^), cg27107882 (P=1.66×10^-03^), and cg11235594 (*NPHP1*, P=6.15×10^-04^). 78.9% of our suggestively significant NeuN+ DMPs had the same direction of effect in the bulk meta-analysis (**Figure 2A**).

**Figure 2:**
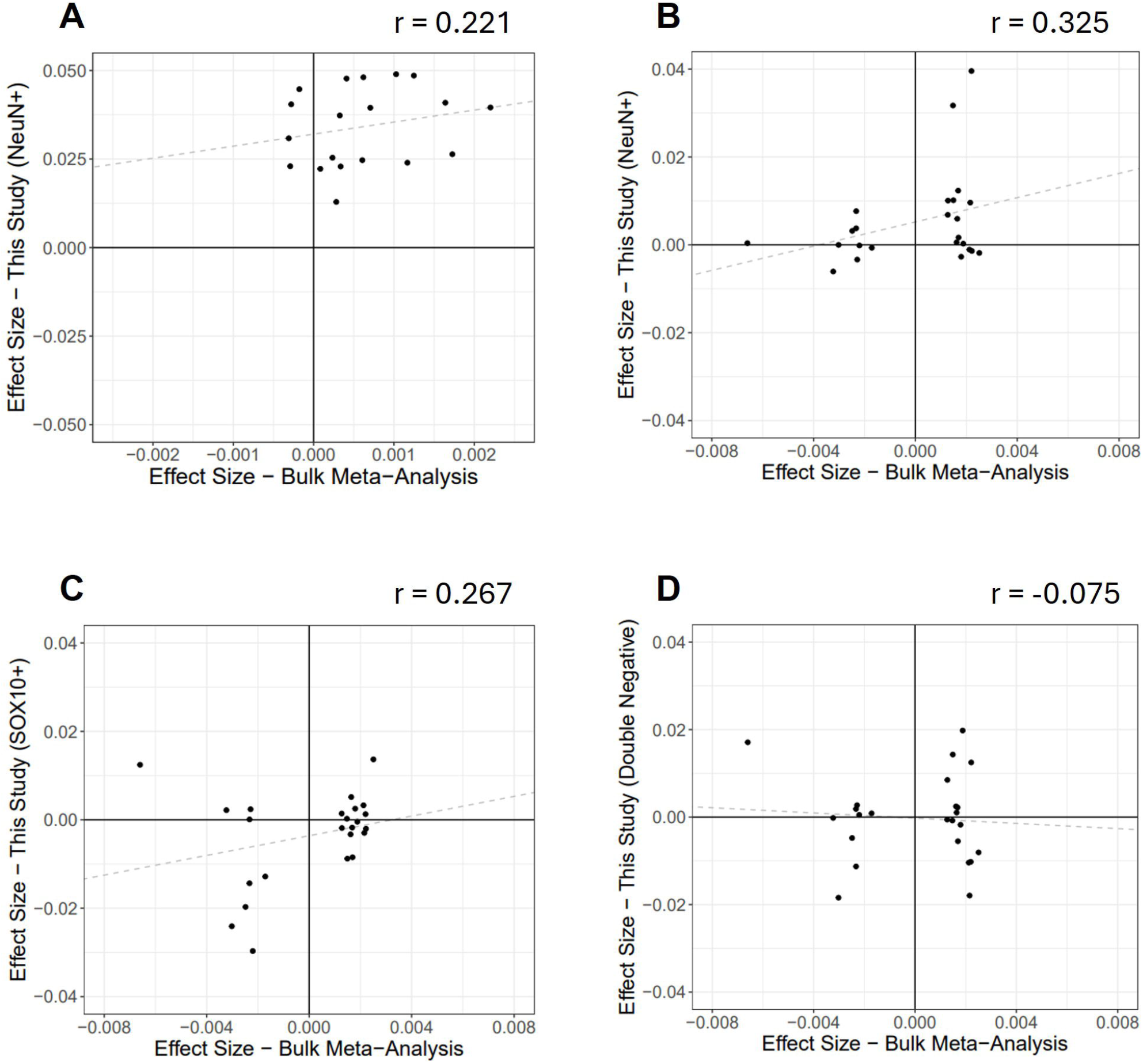
Comparison of EWAS effect sizes with meta-analysis in bulk brain tissue. Comparison of EWAS results with the bulk meta-analysis by Harvey et al. [37]. **(A)** Effect sizes for our suggestively significant NeuN+ DMPs in the bulk data (X-axis) and in our NeuN+ data (Y-axis). Of our 19 sites, 15 sites had a concordant direction of effect between studies (78.9%, binomial test P = 0.019, Pearson correlation coefficient = 0.221). (**B-D**) Effect sizes for the meta-analysis 24 FDR significant DMPs in the bulk data (X-axis) and in our NeuN+ (**B**), SOX10+ (**C**) and double negative (**D**) data (Y-axis). Of their 24 sites, 16 sites had a concordant direction of effect in our NeuN+ data (66.7%, binomial test P = 0.152, Pearson correlation coefficient = 0.325), 12 sites had a concordant direction of effect in our SOX10+ data (50%, binomial test P = 1.0, Pearson correlation coefficient = 0.267), and 11 sites had a concordant direction of effect in our double negative data (45.8%, binomial test P = 0.839, Pearson correlation coefficient = -0.075). For our study, effect sizes are displayed as the mean methylation difference between the PD and control groups, whereas for the bulk meta-analysis effect sizes are displayed as the DNA methylation difference per increasing LB Braak stage. DNA methylation is presented as a proportion. Each dot represents a DNA methylation site. The gray dashed line represents linear regression fit (meta-analysis effect ∼ our effect).

The meta-analysis identified 24 FDR significant DMPs associated with LB stage [37] and one of our suggestively significant PD-associated NeuN+ DMPs (cg13847853) was their top genome-wide significant DMP (P = 2.62E-11). cg13847853 is not annotated to any gene but interestingly the site has been associated with Braak NFT pathology burden [18]. The effects of the 24 meta-analysis DMPs had a greater concordance with our NeuN+ data (66.7% same direction of effect) (**Figure 2B**), compared with our SOX10+ (50% same direction of effect) (**Figure 2C**) and double negative data (45.8% same direction of effect) (**Figure 2D**). This indicates that many of the differences observed in bulk brain tissue may reflect variation in neurons.

## Discussion

To our knowledge, this is the first study in PD to simultaneously profile genome-wide DNA methylation in neurons, oligodendrocytes and other glial cells from the same individuals. Our findings suggest that PD-related DNA methylation differences are enriched in neurons, with the identification of seven genome-wide significant DMPs in neuronal nuclei. These were annotated to *PDE4B* (cg00736744), *SSBP2* (cg09320924), *ROBO4* (cg18248499), *NPHP1* (cg11235594), and *HSD17B12* (cg14489013), as well as two unannotated loci (cg27107882 and cg13471188).

The *PDE4B* gene (encoding phosphodiesterase 4B) is of particular interest in the context of neurodegeneration. Members of the PDE4 family hydrolyze the second messenger cyclic AMP and have been proposed as potential therapeutic targets for PD, AD, and other neurological diseases [57,58]. In experimental PD models, PDE4B inhibition has been shown to reduce α-synuclein levels, prevent MPP^+^/MPTP-induced dopaminergic neuron degeneration, and improve motor performance [59–63]. However, its expression patterns in the human PD brain remain unclear: one magnetic resonance imaging study found elevated PDE4B levels in several brain areas in PD individuals with excessive daytime sleepiness [64], whereas another study observed reduced expression across multiple brain regions, including the cortex, correlating with working memory deficits [65]. Notably, an earlier study that performed bisulfite sequencing in neuronal nuclei from the PD and control cortex identified an enhancer DNA methylation site predicted to target *PDE4B* [46], further supporting the possibility that this gene is epigenetically dysregulated in PD.

As far as we know, the remaining four genes (*SSBP2, ROBO4, NPHP1, HSD17B12)* have not previously been implicated in PD. *SSBP2* encodes single-stranded DNA-binding protein 2, a tumour-suppressor gene that participates in DNA damage response and genome stability. Although its role in PD is unclear, a genomic region containing *SSBP2* has demonstrated a significant inverse causal relationship between educational attainment and AD risk [66]. *ROBO4* encodes the transmembrane receptor roundabout 4, a member of the Roundabout (Robo) family involved in axon guidance. ROBO4 expression is enriched in endothelial cells [67] but its expression has also been observed in the developing rat brain, where it regulated radial cortical neuron migration [68]. *NPHP1* encodes the nephrocystin-1, a protein expressed on microtubule-based structures such as cilia and involved in cell division and adhesion [69]. While not previously linked to PD, certain single nucleotide polymorphisms (SNPs) in the *NPHP1* gene may modify the age of onset in familial AD presenilin 1 mutation-carriers and in late-onset AD [70,71]. *HSD17B12* encodes hydroxysteroid 17-beta dehydrogenase 2, an endoplasmic reticulum–bound enzyme involved in oestrogen metabolism and long-chain fatty acid elongation [72]. This gene has not been previously implicated in any brain-related functions. The remaining two genome-wide significant DMPs (cg13471188 and cg27107882) were not annotated to any gene and, to the best of our knowledge, have not been previously reported in any brain disorder EWAS. GO analysis highlighted that genes annotated to the neuronal DMPs were involved in some processes relevant to the brain and PD. These were photoreceptor connecting cilium (photoreceptor layer thinning, photoreceptor dysfunction and α-synuclein pathology in the retina have been observed in PD [73–76]), transcription coactivator activity (transcription coactivators such as PGC-1α have been strongly implicated in the disease [77,78]), and sensory organ development.

Our power calculations indicated that we had more power to detect differences in the neuronal samples compared to the other cell types. Therefore, the absence of genome-wide significant DMPs in oligodendrocyte and other glial populations may reflect limited statistical power [79]. Given that most complex traits, including PD, are associated with modest effect sizes [56], larger sample sizes will be beneficial in future studies. A limitation of the double negative analysis is the heterogeneity of this population, likely including microglia, astrocytes, and endothelial cells. Refining cell type resolution will be critical for uncovering subtle and cell type–specific DNA methylation changes relevant to PD pathogenesis.

Nearly 80% of our neuronal suggestively significant DMPs had a concordant direction of effect when comparing our study with a meta-analysis previously undertaken in bulk brain tissue by Harvey et al. [37]. The lack of universal overlap could be attributed to the clinical diagnostic differences between the cohorts and different outcome measured in the two studies, with the bulk meta-analysis including a mixture of PD and DLB cases and assessing LB pathology burden rather than PD status. There was a stronger concordance between the bulk DMPs and our neuronal data compared to other cell types, further supporting our main finding of neuronal-specific DNA methylation differences in PD. Interestingly, our findings contrast with those reported in AD, where neuropathology-associated differences were driven by variation in non-neuronal cell types [43]. This may indicate disorder-specific patterns of epigenetic dysregulation affecting distinct primary cell types.

This study represents the first PD EWAS to stratify individuals by *GBA1* status, enabling the investigation of whether DNA methylation at specific sites were influenced by an interaction between *GBA1* status and PD status. We did not find clear evidence of such associations, however sample size was again a limitation, particularly in the control-*GBA1* group. It would be beneficial to have greater-powered studies which stratify PD cohorts by *GBA1* status as well as by other major genetic risk factors to assess whether distinct epigenetic signatures exist across different genetic subtypes of PD.

The work presented here can also be built on by increasing cell type resolution by isolating additional cell populations. Cell type-specific epigenetic changes in other brain regions could also be profiled, such as additional cortical areas, substantia nigra, putamen and cerebellum, all of which are regions where DNA methylation differences have been reported in PD [23,24,28,31,47,50,80]. Additionally, other sequencing platforms such as Oxford Nanopore Technologies can be used to provide full genome-wide coverage and differentiate between methylation and hydroxymethylation [81,82]. Furthermore, integrating DNA methylation data with transcriptomic and proteomic analyses may further clarify the functional consequences of these epigenetic differences and their relevance to PD pathogenesis. Ultimately, this line of research will contribute to the generation of multi-region, cell type–resolved and genotype-stratified epigenomic maps of PD, providing deeper insights into the molecular mechanisms underlying disease heterogeneity and enabling the identification of novel therapeutic targets.

## Materials and Methods

### Human brain samples and bulk DNA extraction

Cryopreserved human post-mortem prefrontal cortex tissue samples were obtained from the UCL Queen Square Institute of Neurology (QSBB) and Newcastle Brain Tissue Resource, which both provided ethical approval for the use of the samples (NHS ethics numbers – 18/LO/0721 for QSBB and 24/NE/0012 for Newcastle). Sample demographic, clinical and neuropathological information (including Braak LB and NFT staging where available) were provided by the brain banks. In total, tissue was obtained from 71 PD individuals (37 PD-*GBA1* and 34 PD-non-*GBA1*) and 56 control individuals who were free of any neurological conditions (9 control-*GBA1* and 47 control-non-*GBA1*). Genomic DNA (used for sample genotype determination) was isolated from ∼25 mg of each sample using the QIAamp DNA Mini Kit (Qiagen), following the manufacturer’s instructions. DNA samples concentrations were measured using the Nanodrop ND-8000 spectrometer. ∼ 50 ng/µl of DNA was required for genome-wide genotyping, with aliquots further diluted for pyrosequencing (∼25 ng/µl).

### Genome-wide SNP profiling and quality control

To obtain genome-wide SNP data, genomic DNA samples from each individual were hybridized to the Infinium Global Screening Array-24 v3.0 BeadChip, and subsequently loaded into the iScan, following the manufacturer’s guidelines. iDAT files containing the raw genotype data were uploaded onto GenomeStudio (v2.0, Illumina) where genotypes were called, using the GRCh37/hg19 manifest. Genomic data QC steps were performed using *PLINK 1*.*9* [83] and according to a standard pipeline in our research group (https://github.com/ejh243/BrainFANS/blob/master/array/SNPArray/preprocessing/1_QC.sh). The Global Screening Array contains probes for several *GBA1* variants and was used to assess the presence of *GBA1* variants in samples.

### Targeted *GBA1* genotyping

Pyrosequencing was used to screen for *GBA1* L444P (rs421016) variants in all samples as this SNP was not present on the genotyping array. Polymerase chain reaction (PCR) forward and reverse and pyrosequencing primers were designed using PyroMark Assay Design software v2. Primers were produced by Integrated DNA Technologies and sequences are shown in **Table 3**. PCR was used to amplify a target region of DNA containing the rs421016 SNP. Each PCR consisted of a 30 µl reaction volume per well: 2 µl DNA and 28 µl master mix. The master mix contained 3 µl 10xBuffer B1, 1.2 µl MgCl_2_ (25 mM), 0.3 µl deoxyribonucleotide triphosphate mix (20 mM), 1.5 µl F/R primer mix (10 µM), 0.3 µl HOT FIRE Polymerase (5 U/µl) and 21.7 µl H_2_O. The PCR stages were as follows: hot start (95°C for 15 minutes), 40 cycles of denaturation (95°C for 30 sec), annealing (62°C for 30 sec) and extension (72°C for 1 min), and a final extension (72°C for 10 min). Following each PCR, 12 samples were selected randomly and run on a 2% SYTO60 gel which was scanned using the Li-COR Odyssey Clx system to confirm the presence of amplified DNA from each sample. rs421016 genotypes were then determined using pyrosequencing, following the manufacturer’s guidelines.

**Table 3:**
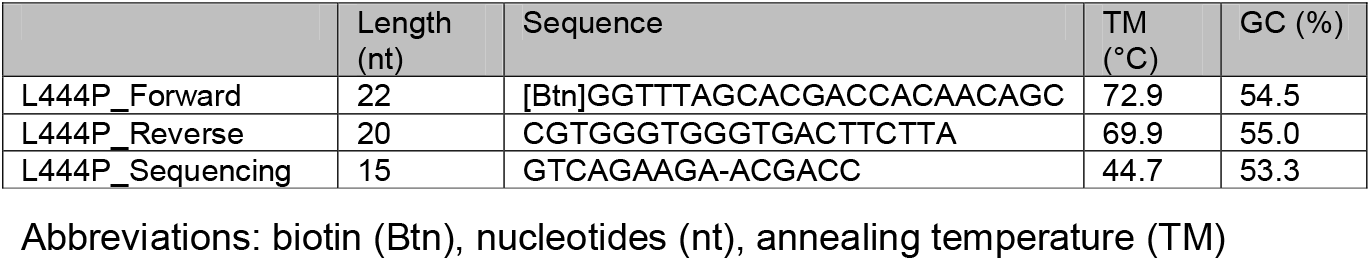
Primer information for L444P genotyping. The L444P PCR forward, PCR reverse and pyrosequencing primer lengths, sequences, predicted annealing temperatures and GC base content, according to the PyroMark Assay Design software.

### Fluorescence-activated nuclei sorting

FANS was performed using a protocol developed in our research group (https://dx.doi.org/10.17504/protocols.io.36wgq4965vk5/v2) [43,52,53], with a few adjustments. ∼250 mg tissue was used as starting material. When resuspending the nuclei pellet with staining buffer prior to immunostaining, the unstained tube and stained tube had a final volume of 750 µl and 400 µl respectively. The following volumes of antibodies were added: NeuN Alexa488 antibody - 1 µl (1:750), SOX10 pre-conjugated antibody - 75 µl (1:10) (QSBB samples) or 25 µl (1:30) (Newcastle samples), IRF8 pre-conjugated antibody - 5 µl (1:150). To decrease costs, the lower SOX10 concentration was used for the Newcastle samples as it resulted in equally effective separation of cell populations. Furthermore, when resuspending the nuclei pellet with staining buffer immediately before sorting, the unstained tube had a volume of 300 µl and the stained tube had a volume of 1 ml. Our FANS gating strategy on a representative sample is shown in **Supplementary Figure 7**. Staining of IRF8 (a microglial nuclei marker) was used to help guide the selection of the double negative population. Up to ∼50,000 NeuN+, SOX10+ and double negative nuclei were collected from each individual. Following nuclei collection, all except the bottom ∼10 µl of supernatant from each tube was removed. Samples were stored at -20°C.

### Bisulfite conversion using DNA Methylation-Direct kit

Sorted nuclei samples (collected in 1.5 ml microcentrifuge tubes) were defrosted at room temperature and then centrifuged at 1,000 x *g* for 1 minute. The EZ-96 DNA Methylation-Direct™ Kit (Shallow-Well Format) (Zymo Research, Cambridge Bioscience, Cambridge UK) was used to bisulfite convert DNA directly from sorted nuclei. To allow samples to fit on 96-well plates, three sorted double negative nuclei fractions from QSBB individuals (with the lowest nuclei numbers) were dropped, leaving a total of 127 NeuN+ samples, 127 SOX10+ samples and 124 double negative samples undergoing bisulfite conversion. The manufacturer’s guidelines were followed, and the eluted DNA was stored at -20°C until the DNA methylation arrays were run.

### DNA methylation profiling and quality control

Prior to running the arrays, the sorted samples within each brain bank were randomized, ensuring there were samples from a mixture of experimental groups and cell types on each array chip, and that the order of experimental groups and cell types varied between array chips. Sorted DNA samples were hybridized to the Infinium MethylationEPIC v1.0 BeadChip (Illumina) and subsequently loaded into the iScan, following the manufacturer’s guidelines (https://support-docs.illumina.com/ARR/Inf_HD_Methylation/Content/ARR/Methylation/Protocol_Manual_fINF_mMeth.htm).

All data analysis was performed using R version 4.2.1. DNA methylation data QC was performed using a standard pipeline built in our research group [79] (https://github.com/ejh243/BrainFANS/tree/master/array/DNAm/preprocessing) which is described below, with sample-level QC output provided in **Supplementary Data 2**.

iDAT files containing the raw signal intensity data were processed using the *bigmelon* package [84]. The median methylated and unmethylated signal intensities were calculated, with all samples passing the exclusion threshold of 500. The *bscon* function (within the *wateRmelon* package) was used to calculate a bisulfite conversion efficiency statistic as the median value across 14 fully methylated control probes. All samples had a bisulfite conversion efficiency > 90%. The percentage of sites with missing data was calculated. Using the *pfilter* function, any samples with detection p-values > 0.05 in more than 1% of probes were removed. In addition, any probes with detection p-values > 0.05 or bead counts < 3 in more than 1% of samples were removed. There were eight SNP probes that were also present on the global screening array which were used to check if the sample genotypes were correlated across the EPIC and SNP arrays, with samples excluded if the correlation was < 0.8. The *predSex* function was used to predict the sex of each sample based on the fold change of the intensity values from probes located on the X and Y chromosomes relative to intensity vales from autosomal probes, with predicted and recorded sex concordant for all samples.

PCA of the autosomal probes was performed using the *prcomp* function from the base stats package in R. This led to the identification of five PCs which explained > 1% of the variance in the raw DNA methylation data (PC1 = 53.4% variance, PC2 = 26.0%, PC3 = 3.6%, PC4 = 2.4%, PC5 = 1.1%). PC1 separated NeuN+ samples from the other fractions, while PC2 separated the SOX10+ and double negative samples from each other. The positions of sorted fractions on PC1 and PC2 were used to assess the accuracy of the cell type isolation, based on guidelines recently published within our research group [79]. This involved the calculation of an average profile for each cell type, based on the mean and standard deviation of PC1 and PC2. A FANS efficiency score was calculated for each individual, showing how similar the sorted samples from a given individual were to expected profiles for each labelled cell type. Individuals who failed the FANS efficiency check (FANS efficiency score > 5) were excluded. Subsequently, any samples which were more than three standard deviations away from all cell type means in either of the first two PCs were also excluded from further analysis.

### Normalization using *adjustedDasen*

All samples remaining after the cell type isolation QC checks were normalized using the *adjustedDasen* function as part of the *wateRmelon* package [85]. Samples in each cell type were normalized separately. The resulting matrix of normalized beta values was used for subsequent analysis.

### CETYGO cell type composition estimation

To provide an additional assessment of the quality of the cell type isolation, the *CETYGO* package (https://github.com/ejh243/CETYGO) [54,55] was used to estimate the cellular composition of all remaining samples. The normalized DNA methylation beta values were used as input and a reference panel consisting of NeuN+, SOX10+ and double negative fractions was selected. Cell type-specific sites were selected by the *CETYGO* package using an analysis of variance (ANOVA) algorithm. For each sample, we used *CETYGO* to estimate the composition of each fraction in the reference panel (composition estimates can slightly exceed 1 due to how they are calculated by the *CETYGO* package). The package also calculates a CETYGO error score for each sample, which is indicative of the accuracy of the cellular composition estimation. The CETYGO error score captures the deviation between a sample’s DNA methylation profile and its expected profile, given the estimated cellular proportions of the sample and the reference profiles of each cell type. Samples were excluded from further analysis if i) the composition estimate corresponding to its labelled cell type was below 0.8, ii) the estimate corresponding to a different cell type to its labelled cell type was greater than 0.2 (**Supplementary Figure 8**), or iii) the CETYGO error score was greater than 0.1. PCA was repeated in the final filtered dataset to visualise how samples were distributed along the first two PCs (**Supplementary Figure 1**).

T-tests were performed (separately in NeuN+, SOX10+ and double negative samples) to determine if any cell composition metrics (calculated using the *CETYGO* package) significantly differed between PD and control samples, considering all remaining samples. The cell composition metrics tested were NeuN+, NeuN-/SOX10+, NeuN-/SOX10-(double negative), NeuN+/SOX6+ (inhibitory GABAergic neuronal enriched), NeuN+/SOX6-(excitatory glutamatergic neuronal enriched), NeuN-/SOX10-/IRF8+ (microglial enriched), NeuN-/SOX10-/IRF8-(triple negative) (astrocyte enriched), and the CETYGO error score.

### Power calculations

Power calculations were performed using the *CellPower* package (https://github.com/ew367/CellPower), which uses the *pwr*.*t*.*test* function from the R *pwr* package [86]. Power calculations were based on a case-control study design and therefore should be interpreted in the context of the case-control analysis. A two-sample t-test was used to compare the means of both groups, where the null hypothesis of each test is that the group means are equal [79]. The effect size (Cohen’s d) is the expected difference between the two group means divided by their pooled standard deviation. This was calculated using the standard deviation of each site for each cell type separately, which was derived from the normalized DNA methylation beta value matrix. Power calculations were based on a genome-wide significance threshold of P = 9.0×10^-08^. Power calculations were performed for each cell type separately based on the smallest group size out of PD and controls after all filtering steps (NeuN+ n=52, SOX10+ n=42, double negative n=45).

### DNA methylation association analyses

For all analyses performed, a series of probes were removed. This consisted of cross-hybridizing and single base extension probes, and probes with a common SNP (European population minor-allele frequency > 0.01) within 10 base pairs of the DNA methylation site [87]. From the initial 815,616 probes, 736,078 probes remained after this filtering step and were included in the EWAS analyses. DNA methylation sites were annotated to genes based on the hg19/GRCh37 annotation built-in to the standard Illumina UCSC gene annotation manifest (using the *IlluminaHumanMethylationEPICanno*.*ilm10b4*.*hg19* package).

First, an EWAS was performed separately in the NeuN+, SOX10+ and double negative samples to identify DMPs associated with PD, regardless of *GBA1* status. A linear regression model was used using the *lm* function in R. The model controlled for the covariates of age, sex and institute (brain bank).

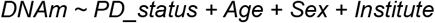

We extracted the effect size (mean difference in DNA methylation beta value in PD compared to controls, where DNA methylation is measured as a proportion), standard error and P value from these regression models. A genome-wide significance threshold of P < 9.0×10^-08^ was used to correct for multiple testing [56], alongside a less stringent suggestive significance threshold of P < 1.0×10^-06^.

To assess the cell type-specificity of the genome-wide significant DMPs, a mixed effects linear regression model was used, which included the data from all three cell types with a random effect to account for the potential correlation between samples from the same individual. The model consisted of main effects for PD status and cell type as well as an interaction term between these (PD_status*Cell_type), to enable all cell types to have their own mean difference between PD and controls estimated. In the model, cell type is represented by two dummy variables – for NeuN and SOX10, where effects were calculated using double negative as the baseline. The model was fitted using the *lmer* function from the *lme4* package. The following regression model was fitted for each DNA methylation site, where i denotes individual and j denotes cell type:

#### Mixed effects full model

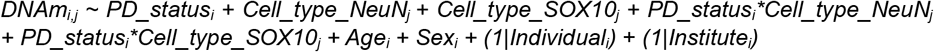

#### Mixed effects null model

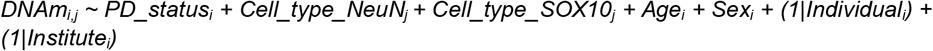

We restricted the analysis to our genome-wide and suggestively significant DMPs (P < 1.0×10^-06^). To assess the cell type-specificity of each DMP, the P value for the interaction term was considered, which was calculated from an ANOVA comparing the mixed effects with an interaction (full model) against a model without the interaction term (null model). A P value threshold of 0.05 was used to determine if the DMP was considered cell type-specific.

To assess whether the DNA methylation levels of any sites were influenced by the interaction between PD status and *GBA1* status, the initial linear regression model was extended to include a term for the *GBA1* status main effect and the interaction between PD status and *GBA1* status. Significant differences in PD effect across *GBA1* subgroups were determined from a t-test of the interaction term. The *lm* function was used, with the following model fitted separately for each cell type:

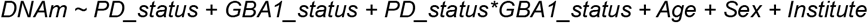

### Gene ontology analysis

We applied GO pathway analysis to the list of genes annotated to the NeuN+ suggestive DMPs (P < 1.0×10^-06^) from the initial case control analysis. DNA methylation sites were annotated to genes using the Illumina UCSC gene annotation. We used logistic regression to test if genes in this list predicted pathway membership, while controlling for gene size (the number of probes annotated to each gene). Pathways were downloaded from the GO website (http://geneontology.org/) and mapped to genes including all parent ontology terms. We considered all genes with at least one probe annotated and mapped to at least one GO pathway. Pathways were filtered to those containing between 10 and 2,000 genes. We applied a Bonferroni significance threshold for the number of pathways tested. The list of significant pathways was further refined by controlling for the effect of overlapping genes. This was achieved by retesting all remaining significant pathways while controlling for the most significant term. If the tested genes no longer predicted the pathway, the term was said to be explained by the more significant pathway, and hence these pathways were grouped together. This algorithm was repeated, taking the next most significant term, until all pathways were considered as the most significant or found to be explained by a more significant term. Finally, we excluded pathways from the results which contained only one gene from the input gene list.

## Supporting information

Supplementary Data 1 FinalSampleNumbers

Supplementary Data 2 QCmetrics

Supplementary Data 3 CETYGOEstimates

Supplementary Data 4 CellCompositionComparisons

Supplementary Data 5 SampleDemographics

Supplementary Data 6 NeuN+LMResults

Supplementary Data 7 SOX10+LMResults

Supplementary Data 8 DoubleNegLMResults

Supplementary Data 9 NeuN+DMPs

Supplementary Data 10 GOAnalysis

Supplementary Data 11 BraakNFTsensitivityAnalysis

Supplementary Data 12 NeuN+InteractionModelResults

Supplementary Data 13 SOX10+InteractionModelResult

Supplementary Data 14 DoubleNegInteractionModelResults

## Data Availability

DNA methylation data will be made available on the Gene Expression Omnibus platform upon final publication.

https://github.com/AnthonyKl98/PD-DNAm-paper

https://github.com/ejh243/BrainFANS/tree/master/array

## Code Availability

DNA methylation quality control scripts used in this manuscript are available at https://github.com/ejh243/BrainFANS/tree/master/array. Scripts used for EWAS analysis are available at https://github.com/AnthonyKl98/PD-DNAm-paper.

## Acknowledgments

We thank researchers for their input including Jonathan Davies, Sam Fletcher, Emma Walker (University of Exeter), and Alison Bernstein (Rutgers University). We acknowledge UCL Queen Square Institute of Neurology and Newcastle Brain Tissue Resource which were used to source tissue samples, along with clinical and neuropathological information used in the study. We also thank the donors who contributed to this study.

## Author Contributions

A.K. conducted laboratory experiments, with methylation array steps carried out by J.B. and genotyping array steps carried out by J.B. and L.W. Laboratory work was supported by B.C., J.B. and A.R.S. A.K. undertook data analysis, with support from E.H. and A.F. E.H. developed the methylation quality control pipeline and analysis models. A.M.R. supported sample selection and obtained samples. J.H., J.I. and K.L. provided additional samples for the research. E.D. developed the FANS method used. A.M.R. and J.M. conceived and supervised the project. A.K. and A.M.R. drafted the manuscript. All authors read and approved the final submission.

## Ethics Declarations

The authors declare no competing interests.

