## Supplementary Figures for "Neuron-Specific DNA Methylation Differences in the Prefrontal Cortex in Parkinson’s Disease"

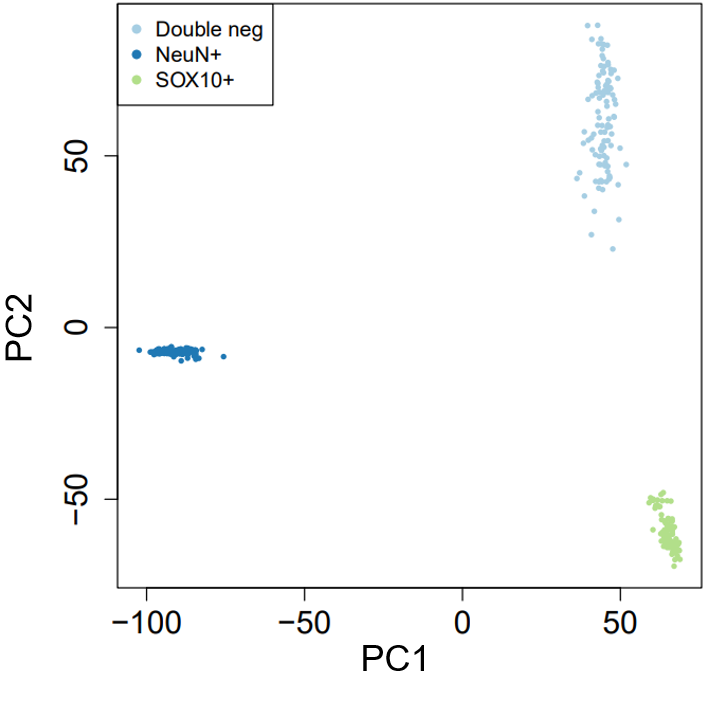

Supplementary Figure 1: Position of samples along the first two principal components of DNA methylation data, color coded by different variables.

Scatterplots of PC1 and PC2, color-coded according to cell type. The final filtered dataset of normalized beta values was used as input. PC1 and PC2 explained 57.2% and 26.9% of variance in the DNA methylation data, respectively. Abbreviation: principal component (PC)

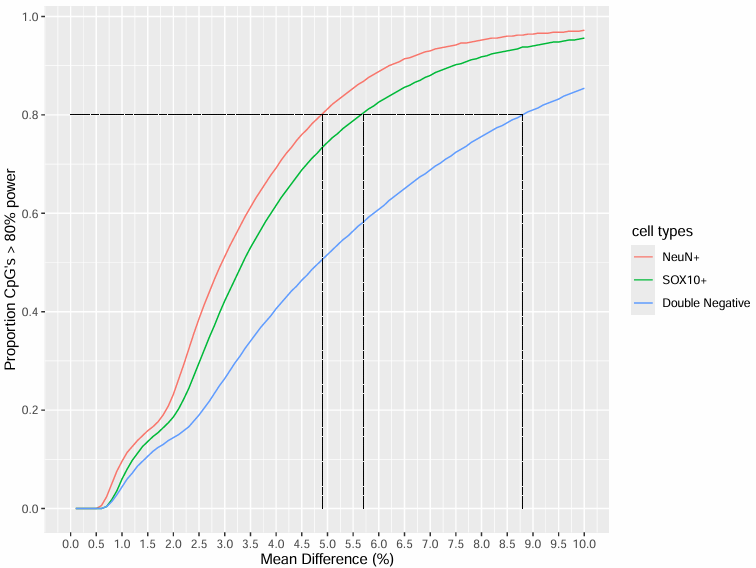

Supplementary Figure 2: Relationship between statistical power and mean difference in DNA methylation between groups for NeuN+, SOX10+ and double negative samples.

The proportion of CpGs with at least 80% power in each cell type for detecting increasing mean differences in DNA methylation (%) between the PD and control groups. Dashed lines denote the mean differences in each cell type that can be detected in 80% of CpGs with > 80% power. Calculations are based on the smallest group size in each cell type remaining after all QC and CETYGO filtering steps, and a genome-wide significance threshold of P < 9.0x10^-08^. Power calculations were performed with the CellPower package, with uses the *pwr.t.test* function. The final normalized DNA methylation beta value matrix was used to calculate the standard deviation of effect sizes for each cell type used in the power calculations.

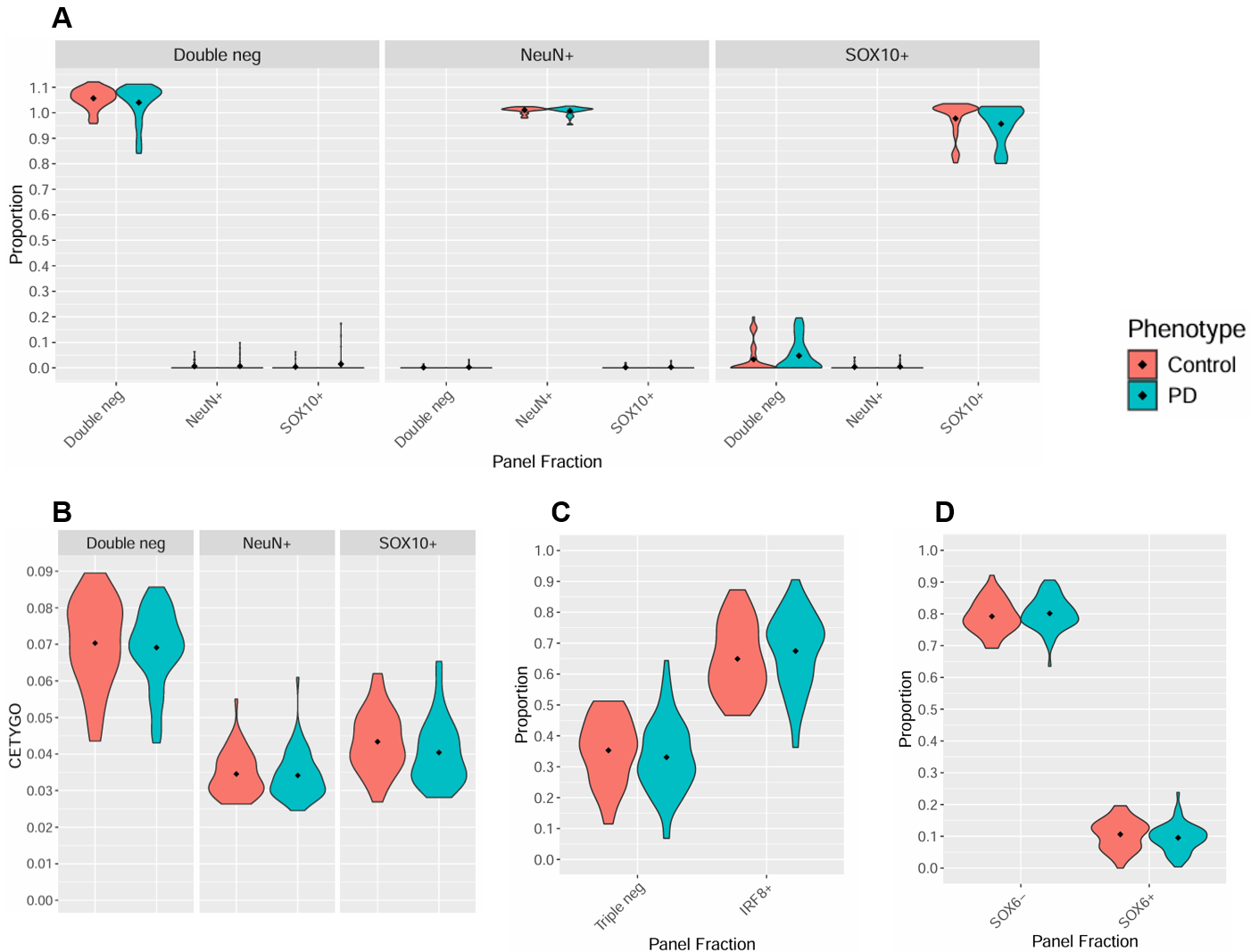

***Supplementary Figure 3: Comparison of cell composition estimates between PD and control samples.***

Cell proportion metric estimates calculated with CETYGO broken down by case-control status. **(A)** The double negative, NeuN+ and SOX10+ proportion scores calculated with CETYGO for our sorted nuclei populations: double negative (left panel), NeuN+ (middle panel) and SOX10+ (right panel). **(B)** The distribution of CETYGO error scores in our sorted double negative (left panel), NeuN+ (middle panel) and SOX10+ (right panel) samples. A higher CETYGO error score indicates a lower degree of accuracy of the cell type composition estimate. **(C)** To assess the contributions of glial cell populations to our double negative nuclei population we used CETYGO to estimate the proportions of microglia (NeuN-/SOX10-/IRF8+) and other glia (NeuN-/SOX10-/IRF8- or “triple negative”) in our double negative samples. **(D)** To assess the contributions of excitatory neurons and inhibitory neurons to our neuronal nuclei population we used CETYGO to estimate the proportions of NeuN+/SOX6- (glutamatergic) and NeuN+/SOX6+ (GABAergic) nuclei in our NeuN+ samples. Abbreviation: cell type deconvolution goodness (CETYGO)

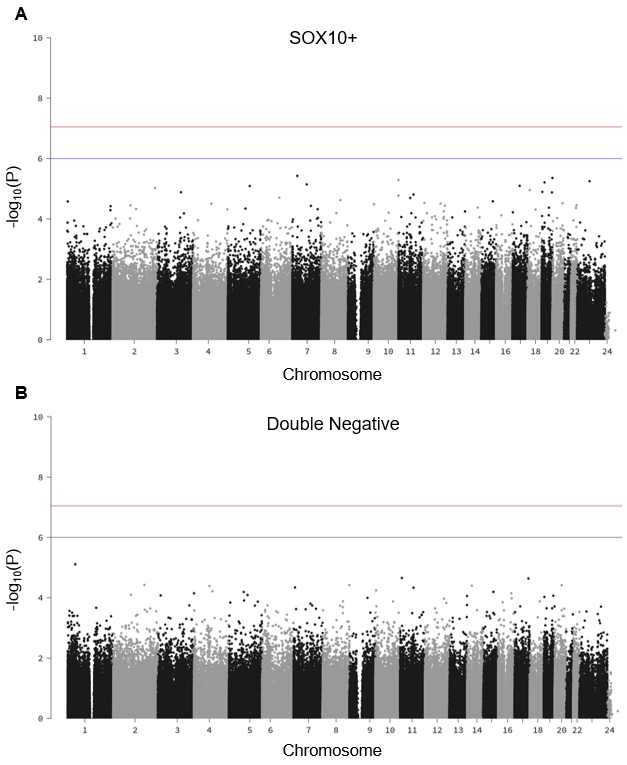

**Supplementary Figure 4: Manhattan plots for EWAS of PD status in the glial populations**

The association of DNA methylation with PD status for (**A**) SOX10+ (oligodendrocyte) (**B**) double negative (other glial) nuclei. The X axis shows chromosome number and the Y axis denotes -log10(P), where a higher value of -log10(P) corresponds to a lower P value. The red line and blue line represent the genome-wide (P = 9.0x10^-08^) and suggestive (P = 1.0x10^-06^) significance thresholds, respectively.

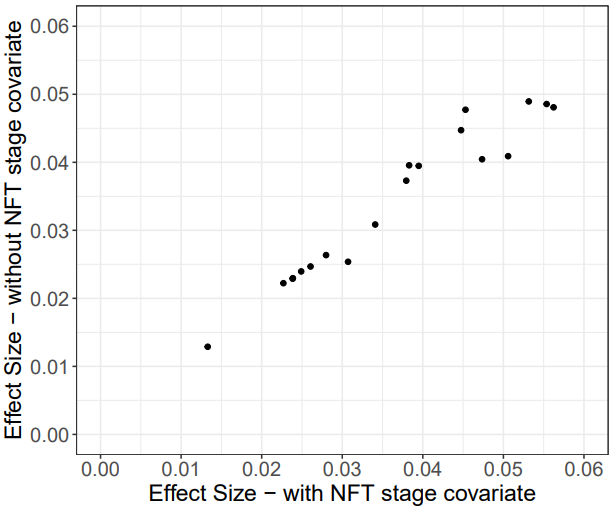

**Supplementary Figure 5: Comparison of NeuN+ EWAS effect sizes with and without adjusting for Braak NFT stage.**

Effect sizes for our suggestive significant NeuN+ DMPs in a sensitivity analysis with Braak NFT stage as an additional covariate (X-axis) compared to the initial EWAS (Y-axis). Effect sizes were highly correlated with all sites showing a concordant direction of effect (Pearson correlation coefficient = 0.968). Effect sizes are displayed as the mean methylation difference between the PD and control groups. DNA methylation is presented as a proportion. Each dot represents a DNA methylation site.

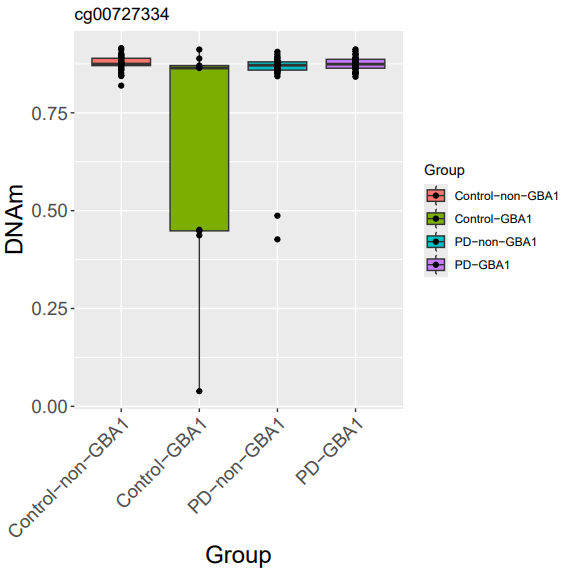

**Supplementary Figure 6: *Distribution of DNA methylation values for cg00727334.***

The distribution of normalized beta values for cg00727334 in control-non-*GBA1*, control-*GBA1*, PD-non-*GBA1*, and PD-*GBA1* groups in NeuN+ samples. Each point denotes an individual. While this DNA methylation site was significantly associated with an interaction between *GBA1* status and PD status, we did not explore this result further due to the presence of outliers. Abbreviation: DNA methylation (DNAm)

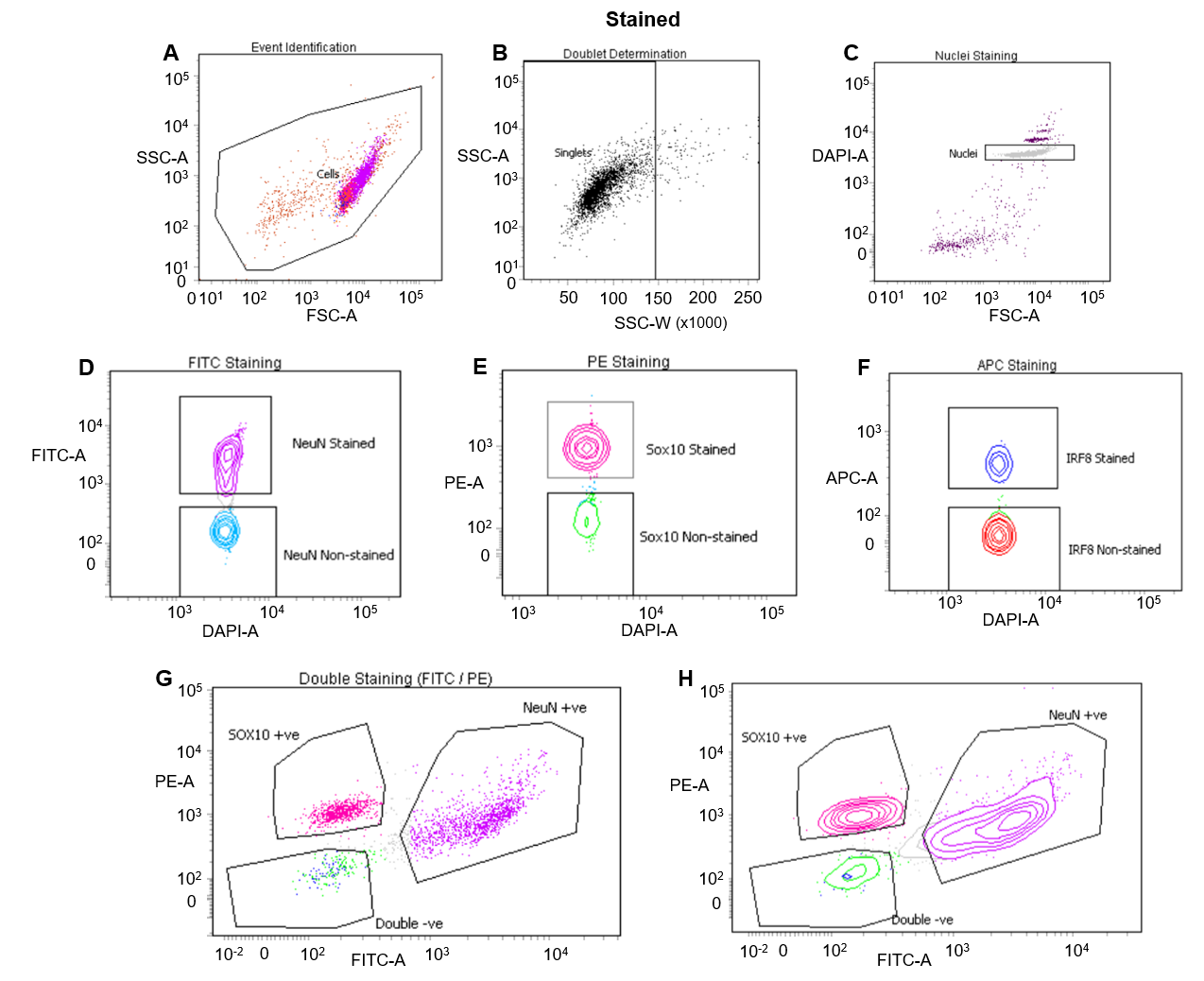

***Supplementary Figure 7****:* ***Fluorescence-activated nuclei sorting gating strategy.***

(**A**) Event Identification: the distribution of events on a plot of FSC-A vs SSC-A. A gate was drawn around the main group of events to be further explored. (**B**) Doublet Determination: on a plot of SSC-W vs SSC-A, a box was drawn around all singlet events, which then excludes any aggregates containing two or more nuclei from being sorted. (**C**) Nuclei Staining: a gate was drawn around the events consisting of intact nuclei which sit in a consistent position on a plot of DAPI-A (nuclei marker) vs FSC-A staining. Any debris or events not consisting of a single nucleus were excluded from being sorted. (**D-F**) Dot and contour plots showing the discrimination of positive and negative populations for each antibody. Note that an unstained sample from the same individual was used as a reference when drawing gates. (**D**) FITC Staining: NeuN-Alexa Fluor488 antibody staining intensity was used to discriminate between neuronal (NeuN stained nuclei) and glial (NeuN non-stained) nuclei. (**E**) PE Staining: considering all NeuN non-stained nuclei, SOX10-NL577-antibody staining intensity was used to discriminate between oligodendrocyte (SOX10 stained) nuclei and other glial (SOX10 non-stained) nuclei. (**F**) APC Staining: considering all SOX10 non-stained nuclei, IRF8-APC antibody staining intensity was used to discriminate between microglia (IRF8 stained) nuclei and remaining other glial (IRF8 non-stained) nuclei. (**G-H**) Stained sample only: final gating on a dot plot (**G**) and contour plot (**H**) of FITC vs PE fluorescence to define the three populations to be collected: a NeuN+ population (magenta) enriched for neuronal nuclei, a NeuN-/SOX10+ (pink) enriched for oligodendrocyte nuclei and a NeuN-/SOX10- (double negative) population (green) enriched for other cell types such as microglia (IRF8 stained nuclei, blue), astrocytes and endothelial cells. The NeuN+, SOX10+, double negative and IRF8 stained events are shown overlaying all events in (**A**). Abbreviations: (FANS), forward scatter (FSC), side scatter (SSC), area (A), width (W)

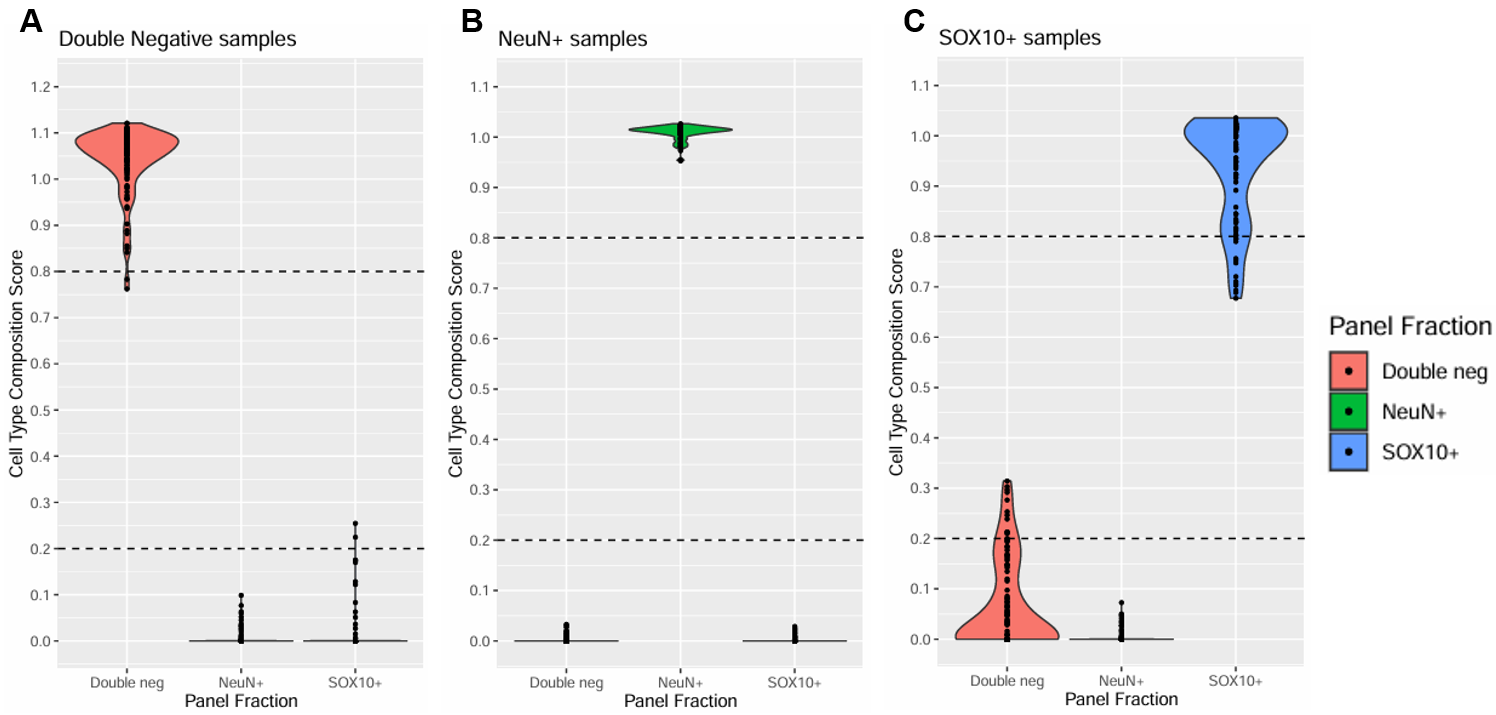

***Supplementary Figure 8: Cellular composition estimates for samples from each sorted population.***

The distribution of double negative, NeuN+ and SOX10+ cell type composition scores (estimated using the CETYGO algorithm and indicative of cell type purity) for (**A**) double negative samples, (**B**) NeuN+ samples and (**C**) SOX10+ samples following initial QC. In each instance, a higher cell type composition score indicates a higher estimate of the corresponding fraction. Samples with a cell type composition score < 0.8 for its labelled cell type or > 0.2 for another cell type (indicated by the dashed lines) were excluded from further analysis.
