## Supplementary Data File Information for "Neuron-Specific DNA Methylation Differences in the Prefrontal Cortex in Parkinson’s Disease"

Supplementary File List

Data 1 File Name: FinalSampleNumbers.xls

Data 1 File Description: Final sample numbers after filtering, grouped by PD and *GBA1* status

Data 2 File Name: QCmetrics.xls

Data 2 File Description: Sample information and QC metrics

Data 3 File Name: CETYGOEstimates.xls

Data 3 File Description: CETYGO cell composition estimates

Data 4 File Name: CellCompositionComparisons.xls

Data 4 File Description: Comparison of cell composition estimates between groups

Data 5 File Name: SampleDemographics.xls

Data 5 File Description: Demographics of samples included in analyses and Tukey HSD test for Braak NFT stage

Data 6 File Name: NeuN+LMResults.xls

Data 6 File Description: Results from EWAS with NeuN+ samples

Data 7 File Name: SOX10+LMResults.xls

Data 7 File Description: Results from EWAS with SOX10+ samples

Data 8 File Name: DoubleNegLMResults.xls

Data 8 File Description: Results from EWAS with double negative samples

Data 9 File Name: NeuN+DMPs.xls

Data 9 File Description: All PD-associated DMPs in NeuN+ samples at P < 1x10^-6^

Data 10 File Name: GOAnalysis.xls

Data 10 File Description: Gene ontology analysis from NeuN+ EWAS

Data 11 File Name: BraakNFTsensitivityAnalysis.xls

Data 11 File Description: Results for 7 NeuN+ DMPs in sensitivity analysis with Braak NFT stage covariate

Data 12 File Name: NeuN+InteractionModelResults.xls

Data 12 File Description: Results from linear regression analysis considering PD and *GBA1* status in NeuN+ samples

Data 13 File Name: SOX10+InteractionModelResults.xls

Data 13 File Description: Results from linear regression analysis considering PD and *GBA1* status in SOX10+ samples

Data 14 File Name: DoubleNegInteractionModelResults.xls

Data 14 File Description: Results from linear regression analysis considering PD and *GBA1* status in double negative samples

Data-file specific information

Data 1 File - FinalSampleNumbers.xls

Variable list:

PD-*GBA1*: PD individuals with *GBA1* variant(s)

PD-non-*GBA1*: PD individuals without a *GBA1* variant

Control-*GBA1*: Control individuals with *GBA1* variants(s)

Control-non-*GBA1*: Control individuals without a *GBA1* variant

NeuN+: Neuronal-enriched nuclei samples

SOX10+: Oligodendrocyte-enriched nuclei samples

Double negative: Other glial cell-enriched nuclei samples

Data 2 File – QCmetrics.xls

Variable list:

Basename: Combination of sentrix ID and sentrix position used to identify sample within lab only

Institute: Brain bank the individual was obtained from

BSConv_Plate: Sample plate during bisulfite conversion

Array_Plate: Sample plate during EPIC array data collection

Array Plate_Location: Sample well during EPIC array data collection

Sex: M/F

Tissue_Type: All prefrontal cortex

Cell_Type: NeuN+ (neuronal), SOX10 + (oligodendrocyte) or Double neg (other glial cells)

Sentrix_ID: EPIC Array ID

Sentrix_Position: Position within EPIC array chip

PD_status: PD or control

GBA1_status: nonGBA or GBA

Dementia_status: Whether the individual was diagnosed with Parkinson’s disease dementia

Braak_LB: Braak Lewy body stage, where information available

Braak_Tangle: Braak tangle stage, where information available

M.median: Median methylated signal intensity

U.median: Median unmethylated signal intensity

intens.ratio: Ratio of median methylated: median unmethylated signal intensity

intensPASS: Pass (TRUE) /fail (FALSE) status of signal intensity check

bisulfCon: Bisulfite conversion efficiency (%)

PC1-6_cp: Values of first six principal components calculated from the control probes of the EPIC array

PC1-5_betas: Values of first five principal components calculated from DNA methylation beta values of the autosomal sites on the EPIC array

pFilter: Pass (TRUE) /fail (FALSE) status of pfilter check

nNAs: The number of sites with missing data

x.cp: Fold change on X-chromosome relative to intensity values from the autosomes

y.cp: Fold change on Y-chromosome relative to intensity values from the autosomes

predSex.x: Predicted sex based on X chromosome fold change

predSex.y: Predicted sex based on Y chromosome fold change

predSex: Overall predicted sex

passFANS: Pass (TRUE) /fail (FALSE) status of FANS efficiency check

maxSD: The number of standard deviations from the cell type mean that the sample is (maximum of PC1 and PC2)

passSDCheck: TRUE/FALSE status that sample falls within required number of SD of cell type means

Other abbreviations used: FANS (fluorescence-activated nuclei sorting), F (female), M (male), PD (Parkinson’s disease)

Data 3 File – CETYGOEstimates.xls

Variable list:

Basename: Combination of sentrix ID and sentrix position

NeuNNeg_SOX10Neg: Double negative (NeuN-/SOX10-) composition estimate

NeuNPos: NeuN+ composition estimate

NeuNNeg_SOX10Pos: SOX10+ (NeuN-/SOX10+) composition estimate

CETYGO: CETYGO error score

Cell_Type: NeuN+ (neuronal), SOX10 + (oligodendrocyte) or Double neg (other glial cells)

PD_GBA1_status: Combination of PD status and *GBA1* status

Passed CETYGO check: PASS/FAIL status of sample after all checks

Other abbreviations used: CETYGO (cell type deconvolution goodness), PD (Parkinson’s disease)

Data 4 File – CellCompositionComparisons.xls

Variable list:

Cell_Type: NeuN+ (neuronal), SOX10 + (oligodendrocyte) or double neg (other glial cells)

Fraction: Reference panel fraction (or CETYGO score) considered

MeanControls: Mean fraction composition estimate (or CETYGO score) in control samples of given cell type

MeanPD: Mean fraction composition estimate (or CETYGO score) in PD samples of given cell type

Pvalue: Assessing if measure significantly differs (P < 0.05) between PD and control groups

Other abbreviations used: CETYGO (cell type deconvolution goodness), PD (Parkinson’s disease)

Data 5 File – SampleDemographics.xls

Variable list:

PD-*GBA1*: PD individuals with *GBA1* variant(s)

PD-non-*GBA1*: PD individuals without a *GBA1* variant

Control-*GBA1*: Control individuals with *GBA1* variants(s)

Control-non-*GBA1*: Control individuals without a *GBA1* variant

P: P value from one-way ANOVA (age, PMI, Braak NFT stage) or χ² test (sex) between PD-*GBA1,* PD-non-*GBA1,* control-*GBA1* and control-non-*GBA1* groups*.*

Mean age (years) (± SD)

Mean PMI (hours) (± SD)

% males

% PDD

Mean Braak LB stage (± SD)

Mean Braak NFT stage (± SD)

Diff: Difference in group means

Lower: Lower bound of 95% confidence interval

Upper: Upper bound of 95% confidence interval

Adjusted P: Adjusted P-value

Other abbreviations used: Honestly significant difference (HSD), Lewy body (LB), neurofibrillary tangle (NFT), Parkinson’s disease dementia (PDD), post-mortem interval (PMI), standard deviation (SD)

Data 6 File – NeuN+LMResults.xls, Data 7 File – SOX10+LMResults.xls, Data 8 File – DoubleNegLMResults.xls

Variable list:

DNAm site

Estimate: Estimate of the mean difference between PD and control groups

SE: Standard error of the estimate

P: P value. Sites where P < 9x10^-08^ are considered genome-wide significant differentially methylated positions, and sites where P < 1x10^-06^ are considered suggestive significant differentially methylated positions.

Other abbreviations used: DNAm (DNA methylation), PD (Parkinson’s disease)

Data 9 File – NeuNDMPs.xls

Variable list:

DNAm site

Estimate: Estimate of the mean difference between PD and control groups in NeuN+ samples

SE: Standard error of the estimate in NeuN+ samples

P (within NeuN+): P value for the estimate in NeuN+ samples

Interaction between PD status and cell type P (considering all cell types): P-value for the interaction between PD status and cell type using a mixed effects model considering all cell types

Chr: Chromosome where site is located

Position: Genomic location of site

UCSC_RefGene_Name: UCSC-annotated gene

UCSC_RefGene_Group: Genomic feature annotation

Relation_to_Island: Relation to CpG island

Other abbreviations used: DNAm (DNA methylation), PD (Parkinson’s disease), TSS (transcription start site), UTR (untranslated region)

Data 10 File – GOAnalysis.xls

Variable list:

GOTerm: Gene ontology term identifier

Name: Name of gene ontology term

Type: Molecular Function (MF), Cellular Component (CC) or Biological Process (BP)

nProbesinPathway: Total number of probes in pathway

nGenesinPathway: Total number of genes in pathway

nTestListProbesinPathway: Number of probes from test list in pathway

nTestListGenesinPathway: Number of genes from test list in pathway

P.GenesinTestList: P value for the gene ontology term

OR: Odds ratio for the enrichment of the GO term in provided gene list

GenesinTestListAndPathway: Lists the genes from test list in pathway

Data 11 File – BraakNFTsensitivity.xls

Variable list:

DNAm site

Estimate: Estimate of the mean difference between PD and control groups

SE: Standard error of the estimate

P: P value for the estimate

Chr: Chromosome where site is located

Position: Genomic location of site

UCSC_RefGene_Name: UCSC-annotated gene

UCSC_RefGene_Group: Genomic feature annotation

Relation_to_Island: Relation to CpG island

Other abbreviations used: DNAm (DNA methylation), PD (Parkinson’s disease), TSS (transcription start site), UTR (untranslated region)

Data 12 File – NeuN+InteractionModelResults.xls, Data 13 File – SOX10+InteractionModelResults.xls, Data 14 File – DoubleNegInteractionModelResults.xls

Variable list:

DNAm site

PD:GBA_Estimate: Estimate of the mean difference in the PD effect between *GBA1* and non-*GBA1* samples

PD:GBA_SE: Standard error of the PD:GBA_Estimate

PD:GBA_P: P value for the PD:GBA_Estimate

PD_Estimate: Estimate of the mean difference in PD compared to controls within non-*GBA1* samples

PD_SE: Standard error of the PD_Estimate

PD_P: P value for the PD_Estimate

GBA_Estimate: Estimate of the mean difference in *GBA1* compared to non-*GBA1* within control samples

GBA_SE: Standard error of the GBA_Estimate

GBA_P: P value for the GBA_Estimate

Other abbreviations used: DNAm (DNA methylation), PD (Parkinson’s disease)
